# Cerebrovascular Reactivity Following Spinal Cord Injury

**DOI:** 10.1101/2022.06.28.22276567

**Authors:** Alexander Mark Weber, Tom E. Nightingale, Michael Jarrett, Amanda H. X. Lee, Olivia Campbell, Matthias Walter, Samuel J.E. Lucas, Aaron Phillips, Alexander Rauscher, Andrei Krassioukov

**Affiliations:** Division of Neurology, Department of Pediatrics, University of British Columbia, Vancouver, BC, Canada; BC Children’s Hospital Research Institute, Vancouver, BC, Canada; School of Biomedical Engineering, University of British Columbia, British Columbia, Canada; Department of Neuroscience, University of British Columbia, Vancouver, BC, Canada; School of Sport, Exercise and Rehabilitation Sciences, University of Birmingham, Birmingham, UK; Centre for Trauma Sciences Research, University of Birmingham, Edgbaston, Birmingham, UK; International Collaboration on Repair Discoveries (ICORD), University of British Columbia, Vancouver, Canada; MRI Research Centre, University of British Columbia, Vancouver, Canada; Department of Urology, University Hospital Basel, University of Basel, Basel, Switzerland; Centre for Human Brain Health, University of Birmingham, United Kingdom; Department of Physiology and Pharmacology, Cumming School of Medicine, University of Calgary, Calgary, Alberta, Canada; Department of Clinical Neurosciences, Hotchkiss Brain Institute, Cumming School of Medicine, University of Calgary, Calgary, Alberta, Canada; Department of Cardiac Sciences, Libin Cardiovascular Institute, Cumming School of Medicine, University of Calgary, Calgary, Alberta, Canada; RestoreNetwork, Hotchkiss Brain Institute, Libin Cardiovascular Institute, McCaig Institute for Bone and Joint Health, Cumming School of Medicine, University of Calgary, Calgary, Alberta, Canada; Department of Astronomy and Physics, University of British Columbia, Vancouver, BC, Canada; G.F. Strong Rehabilitation Centre, Vancouver, BC, Canada; Division of Physical Medicine and Rehabilitation, Faculty of Medicine, UBC, Vancouver, BC, Canada

**Author notes:** denotes equal contribution (Alexander Mark Weber, Tom E. Nightingale). **Address for Correspondence:** A. Krassioukov: ICORD-BSCC, UBC, 818 West 10th Avenue, Vancouver, BC, Canada, V5Z 1M9.

**Keywords:** Spinal cord injuries, blood pressure, tau, steady state cerebrovascular reactivity, functional magnetic resonance imaging

## Abstract

Cervical and upper-thoracic spinal cord injury (SCI) commonly results in autonomic cardiovascular impairments. These impairments can lead to alterations in blood flow, cerebral perfusion pressure and ultimately tissue perfusion, which can lead to an elevated risk of stroke and global cognitive deficits. The aim of this study was to assess cerebrovascular reactivity (CVR) in both the grey matter (GM) and brainstem using functional magnetic resonance imaging (fMRI) in participants with SCI compared to non-injured controls. CVR represents the capacity of brain parenchyma to change cerebral blood flow in response to a vasoactive stimulus (e.g. carbon dioxide, CO_2_) or altered metabolic demand [e.g. neurovascular coupling (NVC)]. Thirteen participants (7 chronic SCI (all male, median age of 42 years), 6 controls (all male, median age of 33 years) were studied cross-sectionally. CVR was measured by assessing the MRI-blood oxygen level–dependent signal with hypercapnic challenge (controlled CO_2_ inhalation). The CVR outcome measure was assessed in three ways. Initially, CVR was calculated as is standard, via the linear, least-squares fit across the whole gas challenge protocol (CVR_whole_). In addition, CVR was further decomposed into its dynamic (tau) and static components (steady state CVR; ssCVR). A 24-hour ambulatory blood pressure monitor was worn to capture free-living blood pressure outcomes. Our results showed a longer tau in the GM of SCI participants compared to controls (median of the difference = 3.0 seconds; p<0.05). Time since injury (TSI) displayed negative correlations with ssCVR in the GM and brainstem of SCI participants: R_S_=-0.77, p=0.041 and R_S_=-0.76, p=0.049, respectively, where R_S_ is the Spearman’s rank Correlation Coefficient. Neurological level of injury (NLI), modified into an ascending, continuous numeric variable, was positively correlated with GM CVR_whole_ (R_S_=0.85, p=0.016), GM ssCVR (R_S_=0.95, p=0.001) and brainstem ssCVR (R_S_=0.90, p=0.006). Lower CVR_whole_ and ssCVR in the SCI-cohort was significantly (P<0.05) correlated with lower daytime blood pressure (R_S_≥ 0.81) and a higher frequency of hypotensive episodes (R_S_≥ -0.83). Thus, living with a SCI for a longer period of time, having a higher NLI and lower blood pressure are linked with poorer CVR outcomes. Our preliminary findings reveal an important difference between the cohorts in the dynamic CVR component, tau. Collectively, these results may partially explain the increased cerebrovascular health burden in individuals with SCI.

**Highlights:**

- CVR is the change in blood flow in response to a vasodilatory stimulus (e.g., hypercapnia).
- Impaired CVR is linked with increased stroke risk and cognitive deficits.
- We investigated the dynamic and steady-state components of CVR using fMRI in individuals with a SCI.
- The dynamic component was significantly different compared to non-injured controls.
- CVR is significantly correlated with time since injury, level of injury and ambulatory daytime blood pressure.

## 1. Introduction

Large population studies have indicated an increased incidence of cerebrovascular disease, such as stroke, in individuals with spinal cord injury [SCI; (Cragg et al., 2013; Wu et al., 2012)]. A recent systematic review also indicated that up to 60% of individuals with SCI suffer from global cognitive deficits (Sachdeva et al., 2018). The cerebrovascular health consequences (i.e., elevated risk-of-stroke and cognitive dysfunction) associated with chronic SCI are likely a result of altered autonomic cardiovascular control (Kim and Tan, 2018; Sachdeva et al., 2019). Indeed, an SCI at or above the sixth thoracic level (≥ T6) can disrupt supraspinal sympathetic control to the heart (preganglionic neurons exiting between T1 - T5 spinal segments) and vascular tone of blood vessels in the trunk and lower extremities (preganglionic neurons exiting between T1 - L2 spinal segments). Consequently, debilitating cardiovascular impairments commonly manifest in these individuals (Phillips and Krassioukov, 2015). These manifestations include resting hypotension, a drop in BP in response to an orthostatic challenge (orthostatic hypotension, OH) and episodes of transient hypertension (autonomic dysreflexia, AD), which is a reflex response to noxious or non-noxious afferent stimuli from below the level of injury, such as a full bladder, constipation, or sexual stimulation. Cerebral autoregulation (CA) ensures that cerebral blood flow is maintained despite changes in perfusion pressure brought about by fluctuations in arterial blood pressure (Willie et al., 2014), although maintained changes in blood pressure can alter cerebral perfusion (Lucas et al., 2010). Moreover, single episodes of extremely high BP can result in acute cerebrovascular consequences (Hubbard et al., 2021; Wan and Krassioukov, 2014), and pre-clinical SCI models have demonstrated structural and functional mal-adaptations in the cerebrovasculature with long-term exposure to repetitive episodes of AD (Phillips et al., 2018; Sachdeva et al., 2020). While chronic hypertension has been highlighted as one of the most prevalent risk factors for stroke (O’Donnell et al., 2010) and vascular cognitive impairment in the general population (van der Flier et al., 2018), less is known regarding the cerebrovascular consequences of transient and repetitive blood pressure fluctuations experienced by humans with SCI.

One major factor shown to predict the risk of ipsilateral stroke (Markus and Cullinane, 2001), and associated with cognitive performance, is impaired cerebrovascular reactivity (CVR) (Catchlove et al., 2018). CVR is now frequently being used as a measure of general brain health, with reduced CVR also being linked to chronic hypertension, dementia and mild cognitive impairment in the general population (Fierstra et al., 2013). CVR represents the capacity of brain parenchyma to change cerebral blood flow in response to a vasoactive stimulus (e.g. carbon dioxide, CO_2_) or altered metabolic demand [e.g. neurovascular coupling (NVC)] (Juttukonda and Donahue, 2019). Previous research has suggested impaired NVC in individuals with chronic SCI, relative to age-matched and sex-matched non-injured controls (Phillips et al., 2014b). A number of studies have looked at CVR responses to a vasoactive stimulus-specifically in people with SCI (Eidelman et al., 1972; Nanda et al., 1976; Wilson et al., 2010). However, conclusions around CVR responses in this population remain equivocal. This is due to different combinations of vasoactive stimuli and neuroimaging modalities being used across these studies, meaning the prior findings are difficult to compare. There is undoubtedly variability in the specificity, sensitivity and spatial and temporal resolution of differing imaging strategies, with a recent study indicating no clear relationship between transcranial Doppler (TCD) and functional magnetic resonance imaging (fMRI) derived CVR measures (Burley et al., 2021). However, the variability of the vasoactive stimulus used in the literature (e.g., hypo/hyper-capnia or hypoxia) or the lack of repeatability in the vasodilatory effect even within a single method, is of a bigger concern (Fierstra et al., 2013). It has been suggested that carbon dioxide (CO_2_) is the most suitable vasoactive stimulus (Fierstra et al., 2013), with increased cerebral blood flow in responses to hypercapnia being mediated by extracellular H+ through H+ sensors expressed on endothelial cells (Wenzel et al., 2020). However, a number of methods can be used to promote hypercapnia, including breath-holding, inspired CO_2_, rebreathing, dynamic end-tidal forcing or prospective end-tidal targeting, each with specific strengths and weaknesses. CVR can now be assessed using a computer-controlled MRI compatible gas delivery system (Fierstra et al., 2013), which provides more precise control over the induced-changes in arterial CO_2_ content and therefore cerebral blood flow.

Perhaps another reason that previous CVR studies have reported equivocal findings in the SCI population is due to the fact that CVR on its own may be too broad of a metric. Indeed, CVR can be further parsed into two metrics: the response time (tau), which is the *rate* of cerebral blood flow (CBF) increase in response to the vasodilatory stimulus; and a static component termed steady-state CVR (ssCVR), which can be thought of as the CVR corrected for the response time (tau). In other words, tau represents the temporal component of CVR and ssCVR represents the time-independent component. In patients with mild dementia and Alzheimer’s disease, for example, CVR response times have been found to be consistently longer than in age-matched participants (Cantin et al., 2011; Cohen et al., 2002; Vazquez et al., 2006). No previous trials in the SCI-population have reported CVR function in terms of both steady state and response times.

To our knowledge, this exploratory study is the first to compare CVR responses following SCI using fMRI with a vasoactive stimulus that is repeatable and reliable. We hypothesized that individuals with SCI would have reduced CVR measures compared to age- and sex-matched non-injured controls, and that these measures would be associated with free-living blood pressure outcomes (i.e., frequency of BP events) and injury characteristics [i.e., time since injury (TSI) and neurological level of injury (NLI)]. Clarifying this question will aid the understanding of the mechanisms underlying changes in cerebrovascular health following SCI and possibly contribute to the development of new rehabilitation therapies in the future.

## 2. Methods

### 2.1 Study Participants

Participants living with SCI were recruited from in and around the Greater Vancouver Area, as were age- and sex-matched able-bodied controls. Detailed inclusion and exclusion criteria are listed in Table 1. All participants provided written informed consent prior to participation in the study, which was approved by the University of British Columbia Clinical Research Ethics Board (H16-01458).

**Table 1:** Participant inclusion and exclusion criteria.

| Inclusion criteria | Exclusion criteria |
| --- | --- |
| *SCI-participants*<br>1. chronic (>1-year post injury) traumatic, motor-complete SCI (AIS A or B) between C4-T6 spinal segments<br>2. documented presence of AD.<br>*All participants*<br>1. Male or female,<br>2. 19 - 65 years of age,<br>3. Willing and able to comply with all clinic visits and study-related procedures. | *All participants*<br>1. History of any of the following medical conditions:<br>i. Untreated cardiovascular disease,<br>ii. Type 1 Diabetes,<br>iii. Untreated Type 2 diabetes,<br>iv. Obesity,<br>v. Severe brain injury,<br>vi. Untreated depression,<br>2. Participants who have a non-MRI safe intrauterine device in place,<br>3. Women intending to become pregnant or were currently pregnant.<br>4. Body mass >158 kg (maximum acceptable weight for the MR scanner),<br>5. claustrophobia,<br>6. severe spasticity,<br>7. Standard exclusion criteria related to MR scanner (e.g. metal implants, cardiac pacemaker, shrapnel) and head circumference >63.5 cm or distance from back of head to tip of nose >24 cm (maximum possible size of a participant's head to fit into the MR head coil with face mask fitted) |

*Abbreviations: AD, autonomic dysreflexia; AIS, American Spinal Injury Association Impairment Scale; C, cervical; MR, magnetic resonance; SCI, spinal cord injury; T, thoracic*

### 2.2 Measuring Cerebrovascular Reactivity

In the brain, the detection of oxygen delivery needs is partly controlled by the presence of CO_2_. Increased partial pressure of CO_2_ (P_a_CO_2_; also known as hypercapnia) will result in a dilation of the cerebral resistance vessels, which in turn results in increased CBF and P_a_O_2_ within the cerebral tissue. Tightly controlled delivery of CO_2_ (and therefore CBF) can be accomplished using a device such as Thornhill’s Respiract Gas Control System (Figure 1A). Given the hypercapnic-induced changes in tissue perfusion, over and above resting metabolism, both arterial and venous oxygenation is elevated during a CO_2_ gas challenge. The resultant change in venous oxygenation can be measured using the blood oxygen level dependent (BOLD) effect seen in fMRI: i.e. the delivery of oxyhemoglobin (HbO) - a diamagnetic molecule - resulting in greater signal detection than deoxyhemoglobin (dHb) - a paramagnetic molecule (Figure 1B). For the purposes of this study, CVR is thus defined and calculated as the percent change in BOLD signal per change in end-tidal partial pressure of CO_2_ (P_ET_CO_2_) and expressed as %ΔBOLD/mmHg.

**Figure 1:**
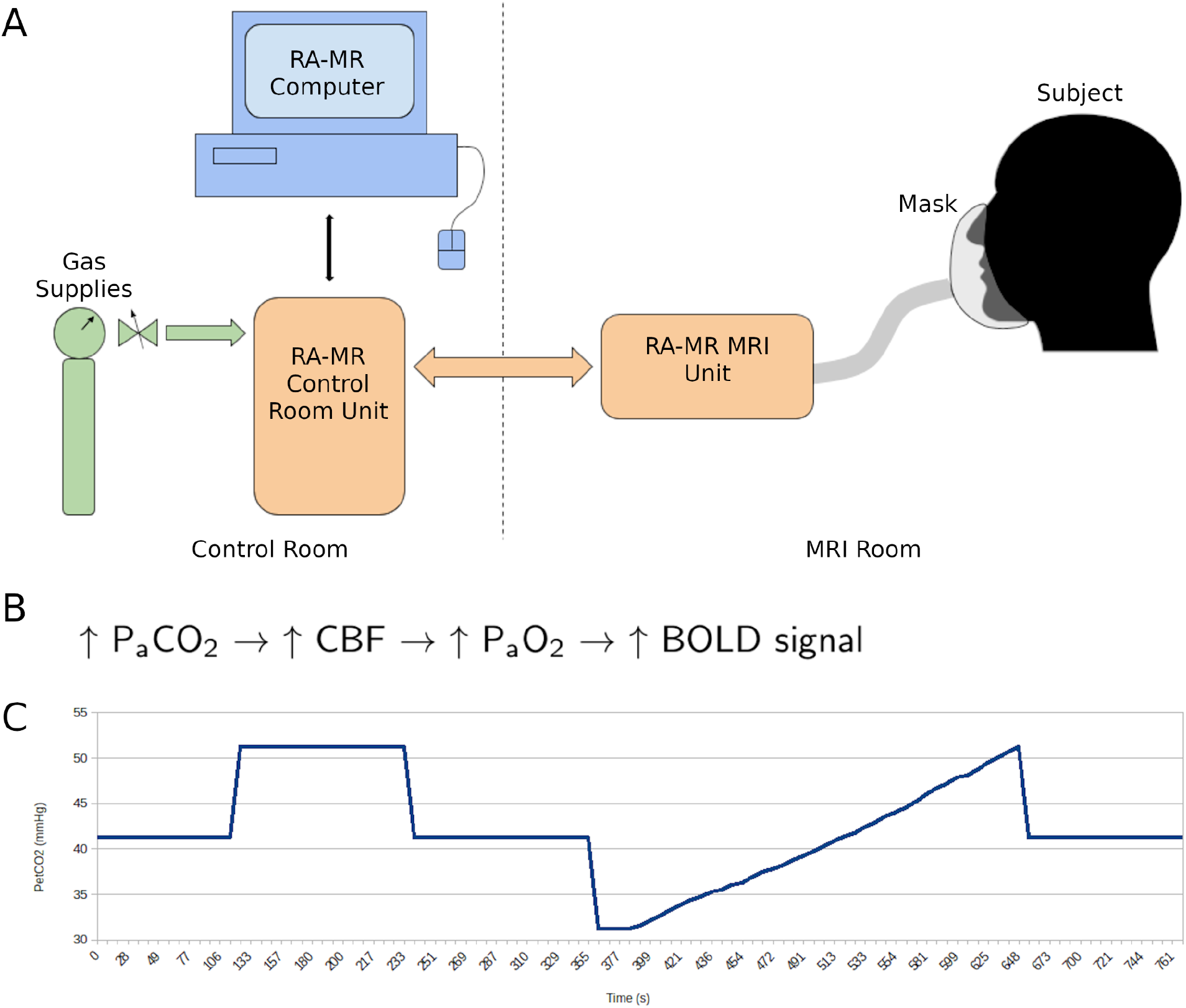
Respiract (RA-MR^TR^) and Cerebrovascular Reactivity (CVR) Schematic Diagram. A) Provides a diagram of the basic components of the RA-MR system. The participant is breathing through a mask that has gas being detected and administered by the RA-MR MRI Unit. This unit in turn receives gas from the RA-MR Control Room Unit, which is connected directly to the gas supplies and the RA-MR Computer. B) The basic sequence of events behind hypercapnia and the increase in fMRI signal. C) Target end-tidal PCO_2_ (P_ET_CO_2_) schematic over the course of the CVR fMRI scan.

### 2.3 MRI Acquisition and Hypercapnic Challenge

All participants were fitted with a new, unopened, and sterile gas mask that was cut to fit each individual’s face. 3M™ Tegaderm™ Transparent Film Dressing (1624W, frame style, 6 cm x 7 cm) were cut in half lengthwise and used to ensure a leak-proof mask fitting. All scans were acquired on a Philips 3T Achieva scanner equipped with Quasar Dual Gradients and an eight-channel SENSE head coil.

The following scans were acquired with the Respiract gas mask on.

Anatomical data were acquired with a high-resolution sagittal 3D T1-weighted turbo field gradient echo (TFE) sequence for registration. The imaging parameters were: repetition time (TR) = 8.1 ms, echo time (TE) = 3.5 ms, flip angle = 8 degrees, acquisition matrix = 256 × 256, field of view = 256 × 256 × 160 mm, isotopic acquired and reconstructed voxel size of 1 mm^3^, SENSE factor of 2 along the left-right direction, scan time = 6m 26s). Functional data were acquired with a 2D echo planar imaging (EPI) gradient-echo fMRI sequence acquired with the hypercapnia challenge (TR = 2,400 ms, TE = 30 ms, flip angle = 85 degrees, acquisition matrix = 64 × 64, field of view = 224 × 224 mm, number of slices = 41, slice thickness = 3.5 mm, slice gap = 0 mm, acquired voxel size = 3.5 × 3.5 × 3.5 mm, reconstructed voxel size = 3.5 × 3.5 × 3.5 mm, EPI factor = 35, 322 volumes, SENSE factor of 2 along the anterior-posterior direction, scan time = 13m 7s). The first four scans were dummy scans to reach steady-state magnetization.

The hypercapnia challenge was achieved by inhalation of CO_2_-enriched air using the computer-controlled gas blender (RespirAct, Thornhill Research Inc., Toronto, ON, Canada). Hypercapnia was employed by applying a target end-tidal PCO_2_ (P_ET_CO_2_) while maintaining end-tidal PO_2_ constant throughout (100 mm Hg). As shown in Figure 1C, targeted P_ET_CO_2_ protocol was delivered as follows: first at baseline (41 mmHg) for 2 minutes, followed by 10 mmHg above baseline (51 mmHg) for 2 minutes, back to baseline for 2 minutes; next, a slow ramp that started with a decrease in 10 mmHg below baseline that proceeds to ramp up to 10 mmHg above baseline for 5 minutes, followed by 2 minutes back at baseline. Heart rate and O_2_ Saturation were acquired on all SCI and a few control participants during the CVR sequence using an Invivo Essential MRI Patient Monitor.

### 2.4 Image Pre-Processing and CVR Calculation

Structural T_1_-weighted image processing included a file conversion to Neuroimaging Informatics Technology Initiative (NIFTI) format, bias-field correction, spatial normalization and segmentation using fmriprep [fmriprep_docker-20.1.3. (Esteban et al., 2017)]; fmriprep uses the FMRIB (Oxford Centre for Functional Magnetic Resonance Imaging of the Brain) Software Library [FSL, (Jenkinson et al., 2012)], Advanced Normalization Tools [ANTs, (Avants et al., 2008)], FreeSurfer (Dale et al., 1999) and Analysis of Functional NeuroImages [AFNI, (Cox, 1996)]. fMRI image preprocessing, including motion correction, slice-timing correction, alignment to T_1_-weighted image, and alignment to Montreal Neurological Institute (MNI) space was performed using fmriprep. CVR maps were calculated as follows. The recorded P_ET_CO_2_ stimulus was time shifted to the point of maximum correlation with the whole brain average BOLD signal. CVR was then calculated as a linear, least-squares fit of the BOLD response (S) to the P_ET_CO_2_ stimulus on a voxel-by-voxel basis across the whole gas challenge protocol (Figure 2). To differentiate this from the other two CVR outcome measures reported, we defined this as CVR_whole_.

**Figure 2:**
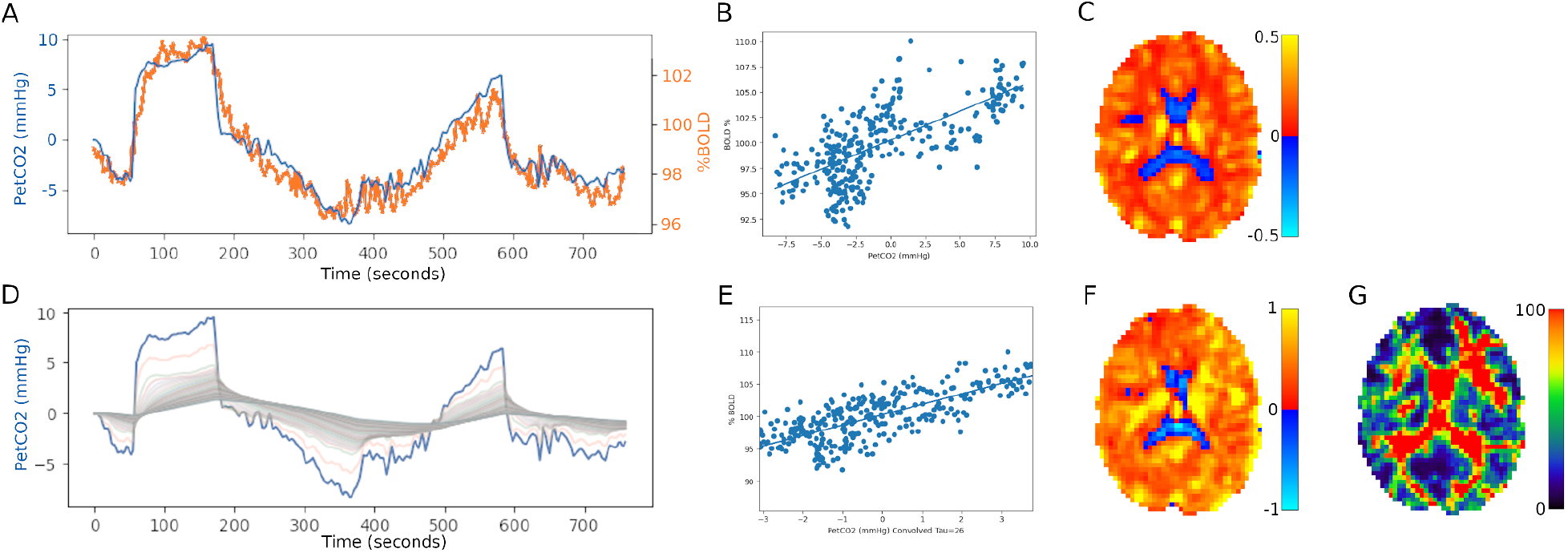
CVR Processing. A) Alignment of P_ET_CO_2_ (blue) with the average BOLD signal (orange). B) Linear least-squares fit of the BOLD response to the P_ET_CO_2_ stimulus on a voxel-by-voxel basis: CVR_whole_. C) Resulting axial slice of a CVR map (in units Δ%BOLD/mm Hg). D) Set of curves resulting from the convolution of P_ET_CO_2_. E) The regression with the best fit between the convolved P_ET_CO_2_ and the BOLD signal: ssCVR. F) Resulting axial slice of a ssCVR map. G) Resulting axial slice of a tau map.

Tau (τ) and steady-state cerebrovascular reactivity (ssCVR) maps were calculated as follows. First the CVR was parsed into a steady-state component (ssCVR) and a dynamic component (τ). The principles of measuring these parameters are outlined schematically in Figure 1 (Poublanc et al., 2015). Briefly, the BOLD response to a stimulus was modeled as the P_ET_CO_2_ convolved with a hemodynamic response functions (HRFs) of the vessels consisting of an exponential decay function of the form: exp(−t/τ), where t is the time variable and τ the time constant of response. Steady-state CVR is calculated by taking the slope of the regression between the convolved P_ET_CO_2_ and Δ%BOLD and can be thought of as the CVR corrected for the speed of the response (τ). In other words, ssCVR represents the time-independent response, with the same units as CVR_whole_ (Δ%BOLD/mmHg), while τ represents the speed of the CVR response to hypercapnia. Regional mean CVR values were obtained from the grey matter (GM) and brainstem masks output as part of the fmriprep analysis.

### 2.5 Free-living Blood Pressure Monitoring in SCI Participants

Ambulatory BP monitoring (ABPM) devices (Meditech ABPM-04, Budapest, Hungary) were used to track the SCI participants’ BP over a continuous 24 hours of daily living. This approach is reliable and well-validated for capturing free-living cardiovascular profiles in individuals with SCI (Hubli and Krassioukov, 2014). BP was measured at 30-minute intervals between 06:00 am and 09:00 am, at 15-minute intervals between 09:00 am and 09:00 pm, and every hour between 09:00 pm and 06:00 am. SCI participants were also instructed to manually record BP and keep a diary to track any key events or record any signs and symptoms of BP instability (i.e. sleep and wake times, bowel movements, bladder emptying, exercise, AD or hypotensive signs and symptoms, etc.) throughout the duration of the 24-hour ABPM assessment day. Baseline BP values were established as the average of three measurements taken from the SCI participants’ nondominant arm while seated in their own wheelchair. These values were subsequently used as a reference point to assess the frequency of AD events during the daytime. AD was characterized as an increase in systolic BP (SBP) >20 mmHg above measured baseline SBP, as per the International Standards to document remaining Autonomic Function after Spinal Cord Injury (Krassioukov et al., 2012). AD events during the nighttime were determined using a baseline nighttime SBP, which was calculated as the average of the first three automatic BP measurements when the participant was asleep. The definition of a hypotensive event is somewhat ambiguous, and in the context of orthostatic hypotension a baseline is often used as the benchmark. We have characterized a hypotensive event using the following criteria that has previously been applied in the SCI-population: SBP ≤100 mm Hg and DBP ≤70 mm Hg (Katzelnick et al., 2019). Hypotensive events were not calculated during the nighttime because of the potential for a nocturnal dip in BP. Average daytime SBP and DBP were also calculated from readings where the participant self-reported as being awake.

### 2.6 Statistical Analysis

All statistics were performed using R (version 4.1.2 - 2021-11-01 - “Bird Hippie”). Due to the small sample size, all statistics were performed and are reported using non-parametric methods. Results are reported using median and interquartile range (IQR) in parentheses. Wilcoxon rank sum tests were conducted to analyze differences between the two cohorts (age, CVR measures, etc.). Spearman correlation were performed for the SCI cohort to explore associations between demographic/injury characteristics (e.g. age, TSI and NLI) and 24-hour ABPM outcomes (e.g. daytime mean SBP and DBP, daily episodes of hypotension and AD), with CVR measures (e.g. CVR_whole_, ssCVR and tau of the GM and brainstem). NLI was modified into an ascending, continuous numeric variable for analysis (i.e. C4 = 1, C5 = 2, C6 = 3 etc.). Significance was set at an alpha level of 5%, uncorrected.

## 3. Results

Ten participants with cervical or upper-thoracic, motor-complete SCI were enrolled in this study. We encountered many obstacles when attempting to send individuals with SCI to the scanner (see limitations). Two participants experienced a panic attack from breathing the CO_2_ rich gas, in which case no fMRI scans were acquired and they were excluded from our analysis. Eventually COVID-19 restrictions and scanner decommissioning put an end to data collection. Because of this, we resorted to using able-bodied participants from the development scans as age- and sex-matched controls. A suitable able-bodied control close in age to ID#2 was not recruited in time and they were excluded from further analysis. Consequently, successful fMRI scans were obtained from seven participants with SCI (all males, with a median age of 42 (16) years and an age range of 21 to 53 years) and six able-bodied controls (all males, with a median age of 33 (8) years and an age range of 24 to 42 years). As all participants were male, no sex differences were explored. No significant difference was found between the age of the two groups (p = 0.283). Table 2 provides a detailed breakdown of individual participant characteristics and scan sequences performed.

**Table 2:** Participant characteristics for individuals with SCI enrolled in the trial and available magnetic resonance scan sequences.

| Participant characteristics |  |  |  |  |  | MRI sequence |  |
| --- | --- | --- | --- | --- | --- | --- | --- |
| ID | Age range [yrs] | Sex | TSI [years] | NLI | AIS | 3DT1 | fMRI (CVR) |
| 1 | 46-50 | M | 2 | T5 | A | ✓ | ✓ |
| 2 | 61-65 | M | 45 | T4 | A | ✓ | ✓ |
| 3 | 31-35 | M | 17 | C5 | B | ✓ | ✓ |
| 4 | 46-50 | F | 34 | T5 | A | ✓ | ✗ |
| 5 | 51-55 | M | 5 | C5 | B | ✓ | ✓ |
| 6 | 31-35 | M | 4 | C6 | B | ✓ | ✗ |
| 7 | 31-35 | M | 13 | T2 | A | ✓ | ✓ |
| 8 | 41-45 | M | 22 | C4 | A | ✓ | ✓ |
| 9 | 21-25 | M | 1 | T3 | A | ✓ | ✓ |
| 10 | 46-50 | M | 22 | C7 | B | ✓ | ✓ |
| All SCI, median (IQR) (n=13) | 44 (17) | 9M/1F | 15 (18) | 5C/5T | 6A/4B |  |  |
| **Participant characteristics for analyzed fMRI scans** |  |  |  |  |  |  |  |
| SCI, median (IQR) (n = 7) | 42 (16) | 7M | 13 (16) | 4C/3T | 5A/2B |  |  |
| Controls, median (IQR) (n = 6) | 33 (9) | 6M | - | - | - |  |  |

Abbreviations: AIS, American Spinal Injury Impairment Scale; ASL, arterial spin labeling; C, cervical; CVR, cerebrovascular reactivity; F, female; fMRI, functional magnetic resonance imaging; M, male; NLI, neurological level of injury; SWI, T, thoracic; TSI, time since injury; 3DT1, 3D T<sub>1</sub>-weighted image

### 3.1 Differences in CVR Metrics Between SCI and Able-bodied Control Participants

Group differences in the GM and brainstem (BS) were explored for CVR_whole_, ssCVR, and tau. A statistically significant difference in tau in the GM between the SCI and able-bodied cohorts was found, with a ∼25% longer tau found in the SCI cohort (median of the difference = 3.00 seconds; p < 0.05). Boxplots are shown in Figure 3.

**Figure 3:**
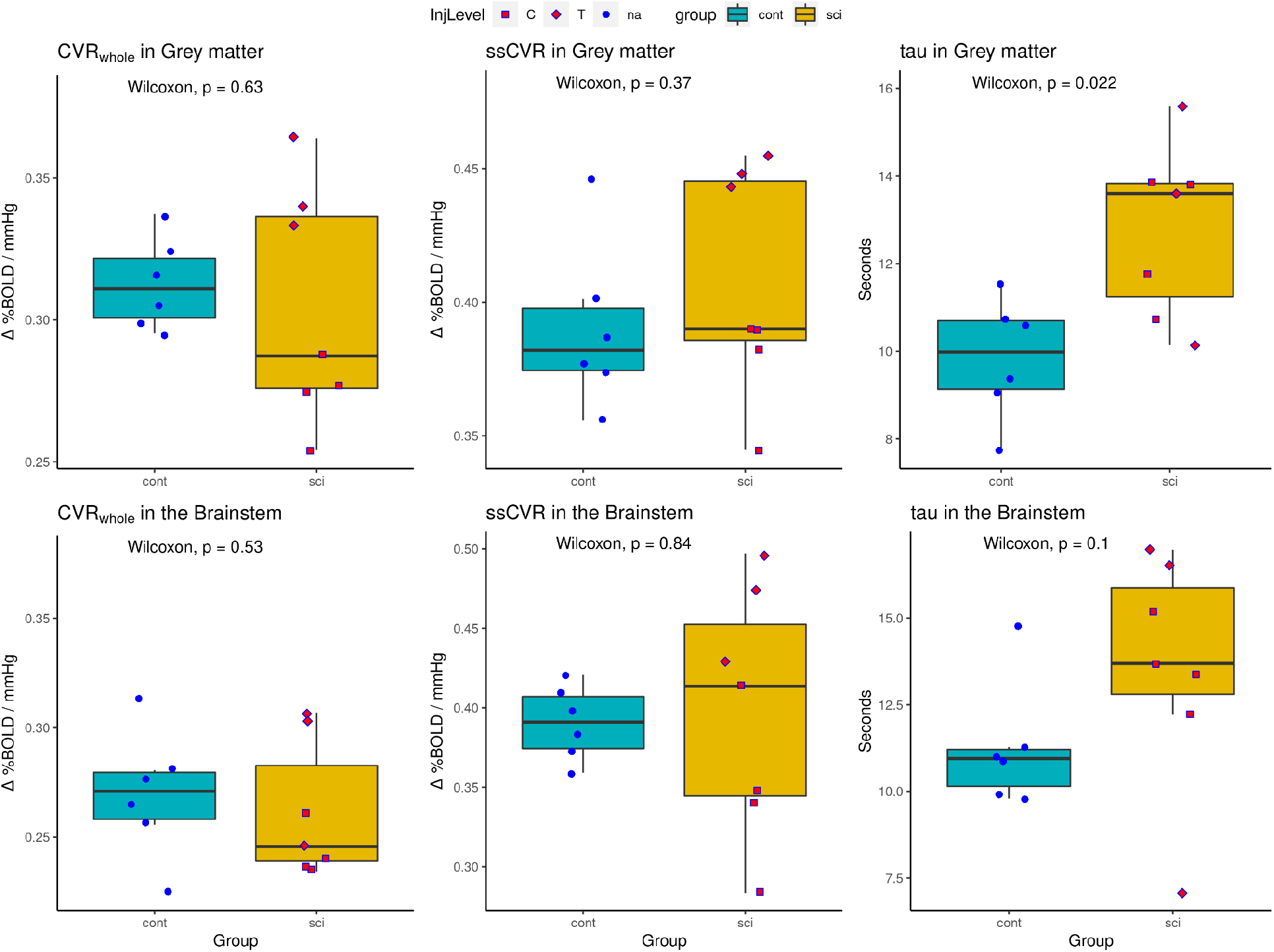
Boxplots Comparing Group Results. Able-bodied controls in cyan color, and SCI participants in yellow. NLI is distinguished as: Cervical = square shape; Thoracic = diamond shape.

### 3.2 Associations Between Injury Characteristics and Free-living BP Variability with fMRI Metrics in the SCI Cohort

As an exploratory analysis, a correlation matrix was created including demographic/injury characteristics (e.g., age, TSI and NLI), physiological outcomes measured during the MR scan [e.g., HR, oxygen saturation (SO_2_)] and 24-hour ABPM outcomes (e.g., mean daytime SBP, DBP and frequency of hypotensive and AD events) with MRI CVR-specific outcomes (e.g., CVR_whole_, ssCVR and tau) in the GM and the brainstem (Table 3).

**Table 3:** Correlation Matrix.

| | Age | TSI | NLI | HR | $SO_2$<br>% | GM<br>$CVR_w$<br>hole | GM<br>ssCV<br>R | GM<br>tau | BS<br>$CVR_w$<br>hole | BS<br>ssCV<br>R | BS<br>tau |
| --- | --- | --- | --- | --- | --- | --- | --- | --- | --- | --- | --- |
| Age |  | 0.13 | -0.18 | 0.23 | **-0.93** | -0.54 | -0.29 | -0.04 | -0.64 | 0.00 | 0.07 |
| TSI |  |  | -0.71 | -0.24 | -0.36 | -0.52 | **-0.77** | 0.63 | -0.59 | **-0.76** | 0.31 |
| NLI |  |  |  | 0.29 | 0.36 | **0.85** | **0.95** | -0.16 | 0.58 | **0.90** | 0.16 |
| HR |  |  |  |  | -0.16 | 0.23 | 0.05 | **0.77** | -0.05 | 0.36 | **0.95** |
| SO<sub>2</sub> % |  |  |  |  |  | 0.58 | 0.40 | -0.11 | 0.75 | 0.18 | -0.05 |
| SBP<br>[n=6] | -0.20 | -0.76 | 0.79 | 0.50 | 0.51 | **0.81** | 0.75 | 0.00 | **0.90** | **0.81** | 0.46 |
| DBP<br>[n=6] | 0.12 | **-0.94** | 0.74 | 0.47 | 0.22 | 0.64 | 0.75 | -0.12 | 0.70 | **0.90** | 0.41 |
| Total<br>AD<br>events<br>[n=6] | 0.60 | 0.20 | 0.12 | 0.20 | -0.62 | -0.14 | 0.14 | -0.09 | -0.71 | -0.03 | -0.09 |
| Total<br>daytim<br>e<br>hypote<br>nsive<br>events<br>[n=6] | 0.09 | 0.52 | **-0.99** | -0.75 | -0.29 | **-0.94** | **-0.94** | -0.20 | -0.60 | **-0.83** | -0.60 |

n=7 unless otherwise noted. Participant ID#11 wore the 24-hr ABPM on two occasions, on both occasions there was an insufficient amount of successful automatic readings (>75%). Values are Spearman Rank correlation coefficients ( $R_s$ ). Bold values have $p < 0.05$ .

CVR_whole_ and ssCVR in the GM and brainstem all showed negative correlations with TSI, with GM ssCVR (R_S_ = -0.77, p = 0.041) and BS ssCVR (R_S_ = -0.76, p = 0.049) being statistically significant. CVR_whole_ and ssCVR in the GM and brainstem were positively correlated with NLI, with GM CVR_whole_ (R_S_ = 0.85, p = 0.016), GM ssCVR (R_S_ = 0.95, p = 0.001), and BS ssCVR (R_S_ = 0.90, p = 0.006) all showing statistically significant correlations. Thus, the more chronic and more severe the injury, the lower ssCVR in the GM and brainstem, and CVR_whole_ in the GM.

CVR response time (tau) in the GM and brainstem were found to have a statistically significant positive correlation with HR (R_S_ = 0.76, p = 0.046 and R_S_ = 0.84, p = 0.019, respectively).

In addition, age and SO_2_ were found to have a strong negative correlation (RS = -0.93; p = 0.003).

The 24-hr ABPM was well tolerated, with a 95% (6%) success rating for automated measurements and total measurements being 66 (11). Daytime SBP and DBP was 107 (15) mmHg and 69 (12) mmHg, respectively, for the included SCI participants (n=6). These participants experienced 6 (13) hypotensive episodes (range: 0 - 53) during the daytime and 8 (5) episodes of AD (range: 2 - 12) over the 24-hr measurement period. In the GM, daytime SBP was statistically significantly correlated with CVR_whole_ (R_S_ = 0.81, p = 0.049), but not ssCVR or tau. In the brainstem, daytime SBP was statistically significantly correlated with CVR_whole_ (R_S_ = 0.90, p = 0.015) and ssCVR (R_S_ = 0.81, p = 0.049), but not tau. Daytime SBP was not found to be statistically significantly correlated with age, HR, TSI, NLI, or SO_2_. In the GM, daytime DBP was not statistically significantly correlated with CVR_whole_, ssCVR or tau. In the brainstem, daytime DBP was statistically significantly correlated with ssCVR (R_S_ = 0.90, p = 0.015), but not CVR_whole_ or tau. Daytime DBP was found to be statistically significantly correlated with TSI (R_S_ = -0.94, p = 0.005) but not with age, HR, NLI, or SO_2_. Total AD episodes were not statistically significantly correlated with any measures. However, hypotensive episodes were found to be negatively correlated with NLI (R_S_ = -0.99, p = 0.0003), CVR_whole_ in the GM (R_S_ = -0.94, p = 0.005), ssCVR in the GM (R_S_ = -0.94, p = 0.005), and ssCVR in the brainstem (R_S_ = -0.83, p = 0.042).

## 4. Discussion

This exploratory study is the first of its kind, in which CVR responses during the delivery of a consistent and repeatable PCO_2_ stimulus were collected using fMRI BOLD and compared between non-injured controls and participants with SCI. As people with SCI face up-to four times the risk of ischemic and hemorrhagic stroke, along with up-to 60% of those injured becoming cognitively impaired (Sachdeva et al., 2018; Wu et al., 2012), it is clear this population is at risk of some form of cerebral health dysfunction. We set out to measure whole-brain cerebrovascular health using multiple CVR measures: CVR_whole_, ssCVR, and tau (CVR response time). The new findings from this preliminary study are that: 1) full cerebrovascular reactivity to CO_2_ (i.e. CVR_whole_) is unchanged in participants with SCI, but this difference may be masked by an increased CVR in thoracic injuries and a decreased CVR in cervical injuries; 2) CVR response times (tau) in participants with SCI were ∼25% higher in GM than in non-injured participants and; 3) CVR response time (tau) in the GM and brainstem were found to have a statistically significant positive correlation with mean HR during the CVR scan. Important injury characteristics linked with the CVR response were identified; 1) ssCVR was found to negatively correlate with TSI in the GM and the brainstem and; 2) CVR_whole_ and ssCVR in the GM and brainstem were positively correlated with NLI, implying improved responses with lower NLI. Lastly, free-living 24-hr ABPM outcomes were associated with the CVR response. Specifically, 1) daytime mean SBP was found to be positively correlated with CVR_whole_ in the GM and CVR_whole_ and ssCVR in the brainstem, 2) DBP was found to be positively correlated with ssCVR in the brainstem, and 3) frequency of hypotensive events were found to be negatively correlated with CVR_whole_, ssCVR in the GM, and ssCVR in the brainstem. Although causation cannot be inferred, collectively, these preliminary associations indicate hypotension (and ultimately brain hypoperfusion) are linked with impaired CVR responses in individuals with SCI.

### 4.1 Previous Brain MRI Studies in SCI

While many studies have performed imaging on the spinal cord itself using MRI (Bejjani et al., 1998; Kuriyama et al., 2009), few have imaged the whole brain (Henderson et al., 2011; Wrigley et al., 2009; Yoon et al., 2013; Zheng et al., 2017). (Zheng et al., 2017) looked at DTI differences between 15 participants with SCI and 15 non-injured controls. They found differences in the white matter in not only the sensorimotor system but also in areas without direct connections to the paralyzed limbs. These changes were also seen to correlate with clinical performance, TSI, and NLI. These findings are evidence that SCI can result in significant broader brain alterations outside the sensorimotor system. With respect to animal models, (Phillips et al., 2018) recently developed a pre-clinical rodent model of AD after SCI. The authors induced repetitive transient hypertension over a four-week period in a group of n=25 rodents after a T3 spinal transection to mimic AD (T3-SCI+AD). Compared to a control group of SCI rats (T3-SCI), the T3-SCI+AD group had impaired cerebrovascular health, including reduced cerebrovascular endothelial function and perivascular sympathetic cerebrovascular innervation. However, they found no differences in CBF between the groups as measured using MRI (ASL) on a 7T scanner. There was also no evidence of cerebrovascular inward remodeling, leading the authors to speculate that repetitive transient hypertension is a unique representation of impaired cerebrovascular health compared to chronic steady-state hypertension in non-injured individuals.

### 4.2 Previous Cerebrovascular Studies in SCI

There have only been a handful of studies that have looked at cerebrovascular function in people with chronic SCI. The results are mixed; with findings showing that high-thoracic and cervical SCI can impair, can have no effect, or can even improve cerebral autoregulation (Kim and Tan, 2018). In 1972, one of the first studies to look at cerebrovascular function after SCI used inhaled ^133^Xe along with hyperventilation (hypocapnia) in 12 participants with cervical injuries (C5-C7) compared to five participants with thoracic (T2-T4) injuries as controls (Eidelman et al., 1972). They found no difference in hypercapnic CBF reactivity between the two cohorts. However, they did find a lack of cerebrovascular responses to hypocapnia in individuals with cervical SCI; with responses in individuals with thoracic injuries being intact. Four years later, using the same technique, (Nanda et al., 1976) found no cerebrovascular response differences to hypocapnia between individuals with cervical injuries (n = 8) and able-bodied controls (and one subject with T2–3 lesion; n = 13). How to reconcile the discrepancy between these two studies remained unclear. Three decades later, (Wilson et al., 2010) used TCD (for blood velocity in middle cerebral artery) and near-infrared spectroscopy (NIRS; for activation-dependent information in prefrontal cortex), to look at hypercapnia and hypocapnia CVR in 6 men with C5-C7 lesions and 14 able-bodied men. They found no difference in either hyper-or hypocapnia between the SCI and able-bodied cohorts. When looking at cerebrovascular resistance (instead of blood velocity or vascular conductance), however, they reported a 30% decrease in SCI participants in response to hypocapnia (but not hypercapnia). While the authors interpreted this result as being reflective of subtle differences in CBF modulation, (Kim and Tan, 2018) in their review of CVR in SCI disagreed, and suggested a more likely explanation could be due to different arterial pressure responses to hypocapnia in individuals with high-level SCI. They reasoned that reduced lung volumes and flow rates due to the disruption of neural innervation of respiratory muscles in SCI leads to imparied mechanics of ventilation, which in turn may impact arterial pressure during hyperventilation. Thus, according to the available literature, whether or not SCI impairs cerebrovascular reactivity remains unanswered and has previously been limited by the assessment methods (e.g. more variability when not using end tidal CO_2_ clamping systems such as the Respiract™ or a lack of spatiotemporal information from TCD) and study designs used (e.g., using individuals with upper-thoracic injuries who may still experience unstable BP as controls).

### 4.3 What Our Preliminary Findings Show

Previous SCI studies have not reported a difference in CVR in response to hypercapnia between SCI and non-injured cohorts. Our study also did not find a difference using CVR on its own (i.e. CVR_whole_). However, by breaking the CVR measure into its steady-state and active components, we reveal an important difference between the cohorts in the response time (tau), and significant correlations between TSI and NLI and the steady state component (ssCVR). These findings may hold the key to explaining the equivocal CVR findings reported in previous studies and highlight the consequences of living with a SCI for a longer time and impact of differing NLI.

#### 4.3.1 CVR_whole_

While no difference in CVR_whole_ was found between the SCI and non-injured cohorts, this may be due to an increased CVR in upper-thoracic thoracic injuries, and a decreased CVR in cervical injuries, in the GM, as can be seen in Figure 3A. Evidence of this sort of overcompensation or adaptation has been reported previously (Wilson et al., 2010). CVR_whole_ was found to be statistically significantly correlated with NLI in the GM. Thus, a lower NLI may have increased CVR compared to non-injured participants, while a higher NLI may have lower CVR compared to non-injured participants. This finding requires replication with a larger cohort.

#### 4.3.2 ssCVR

In all participants, the BOLD signal achieved a better fit to the convolved P_ET_CO_2_ than to the actual P_ET_CO_2_; an example can be seen in Figure 2. This can also be seen in the improved Spearman correlations of TSI and ssCVR in the GM and brainstem (−0.77 and -0.76, respectively) vs. CVR_whole_ (−0.52 and -0.59, respectively). While no differences were found between ssCVR in the two cohorts, ssCVR was found to be significantly correlated with TSI in the GM and brainstem. Thus, the more chronic the injury, the more reduced a participant’s ssCVR. Similar correlations were found when looking at NLI. As we did not find differences in ssCVR between the SCI and non-injured cohorts, this may suggest that acute and lower NLI results in an overcompensation in ssCVR, but that this decreases with time of injury and with higher NLI.

#### 4.3.3 tau

As CVR is by definition a dynamic measure, it logically follows that its speed of response should be examined. A long response time in CVR suggests reduced cerebrovascular health, increased risk of ischemic damage, Alzhimer’s disease or stroke and has been associated with the development of white matter hyperintensities (Duffin et al., 2015; Holmes et al., 2020; Sam et al., 2016). Cerebral autoregulation (CA) is a hallmark of the mammalian brain, and is needed to prevent ischemic damage at low blood pressure (BP) and stroke at high BP (Paulson et al., 1990). These extremes of BP are key characteristics in SCI, especially during OH (Claydon and Krassioukov, 2006) and AD (Krassioukov, 2009). CA adjusts cerebrovascular resistance to ensure that CBF is in line with metabolic needs and is itself composed of both static and dynamic components. Dynamic CA is the rapid regulation of CBF in response to altered arterial BP. Whereas static CA is responsible for limiting CBF changes over gradual perfusion pressure changes. It is interesting that we report differences in the dynamic tau measure (i.e., decreased speed of vascular response) but not ssCVR between the two cohorts. Recent work suggests that the shear-mediated dilation response to a transient CO_2_ stimulus is more endothelium-dependent than the ssCVR component (Hoiland et al., 2022). These data may imply impaired nitric oxide-mediated cerebrovascular function in humans with SCI. Tau was also found to be positively correlated with HR in the SCI cohort, suggesting that the longer tau requires a higher heart rate to respond to a hypercapnic stimulus.

#### 4.3.4 Free-living blood pressure

SBP was found to be positively correlated with CVR_whole_ in the GM and brainstem, and ssCVR in the brainstem. DBP was found to be positively correlated with ssCVR in the brainstem only. Frequency of hypotensive events revealed widespread negative association with NLI, CVR_whole_ (in GM) and ssCVR (in the brainstem). Thus, a lower CVR_whole_/ssCVR was generally correlated with lower blood pressure. It is possible that brain hypoperfusion as a result of low systemic BP is responsible for the lower CVR seen in these participants. These findings are in keeping with other CVR research in the SCI population, albeit when CVR was assessed in response to altered metabolic demand (e.g. NVC) manipulated by an eyes-open and eyes-closed task (Phillips et al., 2014b). Phillips et al., (2014b) demonstrated an absent posterior cerebral artery blood velocity response compared to non-injured controls, and that this was normalized when blood pressure was increased in the SCI cohort using midodrine (an alpha-1 agonist). The authors noted that this improvement in NVC was also related to improved cognitive function. Despite pre-clinical evidence suggesting repetitive exposure to AD results in structural and functional changes in the cerebrovasculature (Phillips et al., 2018; Sachdeva et al., 2020), total frequency of AD events in our study was found not to be correlated with any CVR measure. Looking outside of the cerebrovasculature, (Currie et al., 2019) revealed no correlations with arterial stiffness [measured via carotid-to-femoral pulse wave velocity (cfPWV)] and frequency of AD events derived using 24-hr ABPM, although there were correlations with the frequency of hypotensive events. When AD and hypotensive events (total episodes of BP instability) were combined together significant positive correlations were observed with cfPWV, which is a prognostic risk factor for cardiovascular disease (Vlachopoulos et al., 2010). A recent study raised the possibility that vascular stiffness contributes to altered cerebrovascular hemodynamics and impaired cognitive function in older adults (Bailey et al., 2022). Although our findings suggest a stronger link between low BP and CVR outcomes, the daily fluctuations in BP experienced by this population likely both contribute to the deterioration in cerebrovascular health.

### 4.4 Limitations

Several limitations of the present study should be addressed when interpreting the results. First, the sample size is small with respect to both SCI and non-injured control participants. Recruiting participants with SCI can be challenging (Blight et al., 2019), particularly with specific injury characteristics and hemodynamic instability. This was further compounded when it came to acquiring the necessary medical records detailing spinal hardware or other implants to ensure MR compatibility and safety when scanning at 3T. When MRI compatibility was finally acquired, we encountered further difficulties during scanning. Some participants were unable to fit within the birdcage coil with the Respiract™ mask on, and two participants could not tolerate laying down and breathing the CO_2_ mixture. Lastly, the COVID-19 pandemic and decommissioning of the MR scanner ultimately restricted the recruitment of age and sex-matched, non-injured participants specifically for this study. Other non-injured control participants were used from our development scans and another study that used an identical protocol.

We were unable to measure BP while participants were in the scanner and images were being acquired. An increase in BP has previously been noted during hypercapnia, with less robust increases shown in other clinical groups with disrupted autonomic cardiovascular control (e.g., multiple-system atrophy) compared to controls (Lipp et al., 2010). Not accounting for BP changes during hypercapnia between participants and cohorts means we are unable to provide more insight into the pure ‘cerebrovascular’ response as opposed to perfusion pressure driving the increased CBF. Despite attempts to minimize triggers for AD with gel padding, it is possible that prolonged lying might have resulted in transient elevations in BP in the SCI cohort (Burnham et al., 1994), which may have influenced CVR responses. That being said, heart rate was stable during scans and participants did not exhibit bradycardia, a common physiological sign associated with AD. Body position plays a role in BP and brain perfusion in individuals with cervical and upper-thoracic injuries (Phillips et al., 2014a). Therefore, it remains to be seen whether CVR responses characterized by other imaging modalities [e.g. functional near-infrared spectroscopy (fNIRS), TCD or megnetoencephalography (MEG)] would be more pronounced between individuals with SCI and non-injured controls if the hypercapnic stimulus was provided in the seated position. We wanted to share the aforementioned factors to highlight the logistical difficulties experienced with this study and key considerations for researchers when scanning this population at 3T.

## 5. Conclusion

Our results demonstrate that the CVR response time (tau) to hypercapnia is increased with SCI in GM compared to non-injured participants. Despite decades of equivocal or null findings of hypercapnia CVR in SCI - a mystery given the higher incidence of cerebrovascular health problems in the SCI population - we report for the first time a possible explanation. While CVR_whole_ appears unchanged with SCI, its dynamic and static components reveal another story: 1) that CVR response time is increased (∼25%) in SCI, 2) impaired ssCVR is associated with a greater TSI, and 3) impaired ssCVR is associated with higher NLI’s, who are possibly more predisposed to aberrant blood pressure fluctuations. This assertion is supported by the widespread associations revealed between low blood pressure and CVR outcomes. However, these preliminary findings should be viewed with caution given a lack of corresponding BP data collected during that MR scan, small sample size and cross-sectional study design. Further work is necessary to support these observations.

## Data Availability

All data produced in the present study are available upon reasonable request to the authors

## Acknowledgments

We would like to thank the research coordinators [Andrea L. Ramirez, Martina Franz, Tahira Tejpar and Laura McKracken (all ICORD)] working on the trial between 2016 - 2022 and MRI technicians (Laura Barlow, Neil Wiley, Rick West, and Alex Mazur, UBC) for their assistance with screening and recruiting participants and acquiring scans, respectively. We would also like to thank Megan Ormond (University of Birmingham) for helping to process the 24-hr ABPM data and the participants for their time and commitment.

## Funding

Alexander Weber was a recipient of a Child & Family Research Institute M.I.N.D. Postdoctoral Fellowship from BC Children’s Research Institute, Vancouver, BC. Tom Nightingale (grant number 17767) and Matthias Walter (grant number 17110) were recipients of Michael Smith Foundation for Health Research Trainee Awards in conjunction with the International Collaboration on Repair Discoveries and Rick Hansen Foundation, respectively. The Phillips Lab is supported by the Wings for Life Foundation (Project Grant), Compute Canada (Resources for Research Groups), Natural Sciences and Engineering Research Council (Canada; Discovery Grant), the Canadian Institutes of Health Research (Project Grant), Alberta Innovates Health Solutions, Campus Alberta Neuroscience, the Libin Cardiovascular Institute of Alberta, the Hotchkiss Brain Institute, and the Rick Hansen Institute. Dr. Rauscher is supported by Canada Research Chairs. Dr. Krassioukov’s laboratory is supported by funds from the Canadian Institutes for Health Research, Heart and Stroke Foundation, Canadian Foundation for Innovation, BC Knowledge Development Fund, and Craig H. Neilsen Foundation. Dr. Krassioukov is supported by an Endowed Chair, Department of Medicine, University of British Columbia. The study was funded by a Heart & Stroke Canada Foundation grant (#G-16-00012571. PI: Dr. Krassioukov).

## Declarations of Interest

none

## Notes

### Competing Interest Statement

The authors have declared no competing interest.

### Author Declarations

All participants provided written informed consent prior to participation in the study, which was approved by the University of British Columbia Clinical Research Ethics Board (H16-01458).

